# An Introduction to Proximal Causal Learning

**DOI:** 10.1101/2020.09.21.20198762

**Authors:** Eric J Tchetgen Tchetgen, Andrew Ying, Yifan Cui, Xu Shi, Wang Miao

## Abstract

A standard assumption for causal inference from observational data is that one has measured a sufficiently rich set of covariates to ensure that within covariate strata, subjects are exchangeable across observed treatment values. Skepticism about the exchangeability assumption in observational studies is often warranted because it hinges on investigators’ ability to accurately measure covariates capturing all potential sources of confounding. Realistically, confounding mechanisms can rarely if ever, be learned with certainty from measured covariates. One can therefore only ever hope that covariate measurements are at best proxies of true underlying confounding mechanisms operating in an observational study, thus invalidating causal claims made on basis of standard exchangeability conditions. Causal learning from proxies is a challenging inverse problem which has to date remained unresolved. In this paper, we introduce a formal potential outcome framework for *proximal causal learning*, which while explicitly acknowledging covariate measurements as imperfect proxies of confounding mechanisms, offers an opportunity to learn about causal effects in settings where exchangeability on the basis of measured covariates fails. Sufficient conditions for nonparametric identification are given, leading to the *proximal g-formula* and corresponding *proximal g-computation algorithm* for estimation. These may be viewed as generalizations of Robins’ foundational g-formula and g-computation algorithm, which account explicitly for bias due to unmeasured confounding. Both point treatment and time-varying treatment settings are considered, and an application of proximal g-computation of causal effects is given for illustration.

## 1 Introduction

A key assumption routinely made for causal inference from observational data is that one has measured a sufficiently rich set of covariates, to ensure that within covariate strata, subjects are exchangeable across observed treatment values^1,2^. This fundamental assumption is inherently untestable empirically, without introducing a different untestable assumption, and therefore must be taken on faith even with substantial subject matter knowledge at hand. For this reason, the assumption of exchangeability in observational studies is often the subject of much skepticism, mainly because it hinges on an assumed ability of the investigator to accurately measure covariates relevant to the various confounding mechanisms potentially present in a given observational study. Realistically, confounding mechanisms can rarely if ever, be learned with certainty from measured covariates. Therefore, the most one can hope for in practice, is that covariate measurements are at best proxies of the true underlying confounding mechanism operating in a given observational study. Such acknowledgement invalidates any causal claim made on the basis of exchangeability. In this paper, we introduce a general framework for *proximal causal learning*, which while explicitly acknowledging covariate measurements as imperfect proxies of confounding mechanisms, enables one to potentially learn about causal effects in settings where exchangeability does not hold on the basis of measured covariates.

As all formal methods for causal inference, the proposed proximal approach relies on assumptions that are not testable empirically without a different assumption; nevertheless, as we argue next, we view the required identifying assumptions as easily interpretable and potentially easier to reason about on subject matter grounds than exchangeability. Mainly, proximal causal learning requires that the analyst can correctly classify proxies into three bucket types: a. variables which are common causes of the treatment and outcome variables; b. treatment-inducing confounding proxies versus c. outcome-inducing confounding proxies. A proxy of type b is a potential cause of the treatment which is related with the outcome only through an unmeasured common cause for which the variable is a proxy; while a proxy of type c is a potential cause of the outcome which is related with the treatment only through an unmeasured common cause for which the variable is a proxy. Proxies that are neither causes of treatment or outcome variable may belong to either bucket type b or c. Examples of proxies of type b and c abound in observational studies. For instance, in an observational study evaluating the effects of a treatment on disease progression, one is typically concerned that patients either self-select or are selected by their physician to take the treatment based on prognostic factors for the outcome; therefore there may be two distinct processes contributing to a subject’s propensity to be treated. In an effort to account for these sources of confounding, a diligent investigator would endeavor to record lab measurements and other clinically relevant covariate data available to the physician and the patient when considering treatment options. For instance, it is customary in evaluating the effectiveness of HIV anti-retroviral therapy, to adjust for CD4 count measurement as a potential source of confounding by indication^3^; this is because whenever available to the prescribing physician, CD4 count measurement is invariably used to decide (at least prior to the advent of universal test and treat) whether a patient should be given ART, i.e. the probability of treatment initiation generally decreases with a patient’s increasing CD4 count. However, as an error-prone snapshot measurement of the evolving state of the patient’s underlying immune system, CD4 count measurement is unlikely to be a direct cause of disease progression but rather a proxy of the actual state of patient’s immune system, the actual cause of disease progression. It is therefore more accurate to conceive of baseline CD4 count measurement as an imperfect proxy of immune system status. In addition, two patients with similar CD4 count measurements seen by different physicians may differ in terms of treatment decisions depending on the patient’s own health seeking behavior, as well as differences in physician’s clinical training and experience, and overall prescription preferences; which are all factors potentially predictive of patient disease progression regardless of treatment. Because such factors are notoriously difficult to measure accurately, they are likely to induce residual confounding, even after adjusting for baseline CD4 count.

A proxy of type c might include baseline covariate measurements assessing a patient’s mental and physical commorbidities including those measured using a validated questionnaire. For instance, it is well known that in addition to the immune system, HIV affects the nervous system and the brain producing neurological sequelae, often resulting in forgetfulness and cognitive problems^4,5^. These problems can compromise medication adherence, interfere with instrumental activities of daily living such as driving and managing finances, increase dependency, and decrease quality of life. Several cognitive functioning screening tools exist to objectively measure cognition, with the gold standard being the Mini-Mental State Examination (MMSE)^6.7^, which is a 30-point questionnaire that is used extensively in clinical and research settings. A score of 24 or less is generally used as a cut-off to indicate possible mild cognitive impairment or early stage dementia^6^ although the cut-off can vary according to the education level of the individual^8^. Although widely used as a measure of cognition, MMSE is at best an imperfect proxy of a patient’s baseline state of cognitive impairment, which may in turn influence both a patient’s willingness to initiate and adhere to ART, and the patient’s disease progression at follow-up. Thus, in evaluating the causal effect of ART on disease progression such as say cognitive decline, baseline MMSE measurements can be seen as a proxy of type c for the underlying confounding mechanism corresponding to the patient’s underlying state of cognitive impairment at baseline. These are but two motivating examples of confounding proxies in analysis of the causal effects of ART on HIV infection related disease progression from an observational study. Aside for these proxies, there may also be factors that can accurately be described as true common causes of treatment and outcome processes; these variables which we have referred to as of type a may in fact include age, gender and years of education depending on the context. Thus, rather than as current practice dictates, assuming that adjusting for baseline covariates, exchangeability can be attained, our proposed proximal framework requires the investigator correctly classifies covariates that belong in bucket types a, b and c without necessarily the need for exchangeability to hold conditional on such proxies.

In order to ground ideas, we briefly describe the proposed proximal approach in the context of a point exposure *A*, outcome *Y*, and unmeasured confounder *U*; then suppose that one can correctly select a treatment-inducing proxy *Z* and an outcome-inducing proxy *W* such that the simple structural linear model given below holds:

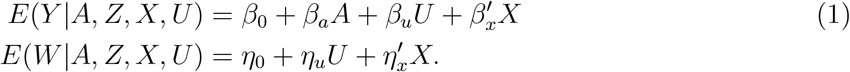

where *X* are all other observed covariates, and validity of proxies is encoded by the fact that the right handside of the first equation does not depend on *Z*, the right handside of the second equation does not depend on *A* and *Z*; and *W* is *U* relevant in the sense that *η*_*u*_ ≠ 0. The causal parameter of interest is *β*_*a*_ = *E*(*Y*_*a*+1_ −*Y*_*a*_| *U, X*) corresponding to the average outcome difference if one were to intervene to increase the treatment by one unit upon conditioning on covariates (*U, X*) a sufficient confounding adjustment set; that is exchangeability holds conditional on (*U, X*). It is then straightforward to show that

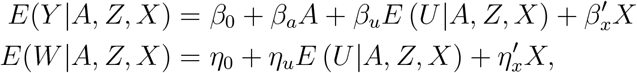

so that

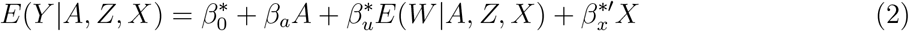

where

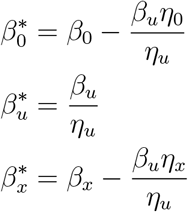

Let 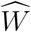 denote an (asymptotically) unbiased estimator of *E*(*W* |*A, Z, X*), then equation (2) suggests that provided that *E* (*U* | *A, Z, X*) depends on *Z*, the least squares linear regression of *Y* on 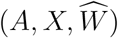 recovers a slope coefficient for 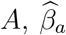 that is consistent for the causal parameter *β*_*a*_. In contrast, either removing 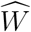 from the regression model, or replacing it with either *W* or (*W, Z*) will generally yield a biased estimate of *β*_*a*_ given that exchangeability does not hold either conditional on *X*, on (*X, W*), or on (*X, W, Z*). As further adjusting for 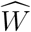 debiases the least squares estimator of *β*_*a*_ conditional on *X*, we shall refer to 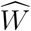 as a *proximal control variable*. The system of linear structural equations considered above is overly restrictive, assuming linearity and no interactions; as we demonstrate in this paper, these assumptions are not strictly necessary and can be relaxed considerably so that nonparametric identification remains possible under certain conditions.

Notation and formal definitions used throughout the paper are given in the next section, non-parametric identification conditions for proximal causal learning are presented in the following section, where it is shown that causal effects can sometimes be identified by the *proximal g-formula*, a generalization of Robins’ foundational g-formula which accounts for confounding bias due to unmeasured factors. For estimation, a *proximal g-computation algorithm* is then introduced. As we show, equation (2) can be recovered as a special case of proximal g-computation algorithm. Both point treatment and time-varying treatment settings are considered, and applications of proximal causal learning are given to illustrate the methodology. The paper concludes with brief final remarks.

## 2 Notation and definitions

Suppose one has observed i.i.d samples on (*A, L, Y*) where *A* denotes a treatment of interest, *Y* is an outcome of interest and *L* is a set of measured covariates. Let *Y*_*a*_ denote the potential outcome had, possibly contrary to fact a person received treatment *A* = *a*. Throughout, we make the standard consistency assumption linking observed and potential outcomes

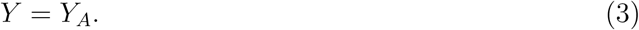

We aim to identify a population average causal effect, corresponding to a contrast of counterfactual averages *β* (*a*) = *E*(*Y*_*a*_) for different values of *a*. For instance, in case of binary treatment, one might be interested in the average treatment effect measured on the additive scale *β* (1) − *β* (0) = *E*(*Y*_1_) − *E*(*Y*_0_); in case of binary outcome, one might also be interested in the average treatment effect on the multiplicative scale *β* (1) − *β* (0) = Pr(*Y*_1_ = 1)/Pr(*Y*_0_ = 1). In all cases, whether *A* is binary, polytomous or continuous, learning about causal effects on any given scale involves learning about the potential outcome mean *β* (*a*), which we aim to identify from the observed sample. A common identification strategy in observational studies is that of exchangeability^1,2,9^ or no unmeasured confounding (NUC) condition on the basis of measured covariates:

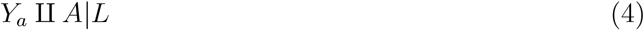

where ⨿ denotes independence, together with positivity condition that

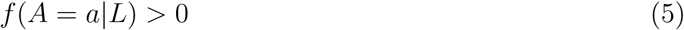

where *f* (*d* | *g*) denotes the conditional density or probability mass function of *d* given *g*. Assumption (4) is sometimes interpreted as stating that *L* includes all common causes of *A* and *Y*; an assumption represented in causal directed acyclic graph (DAG) in Figure 1.a. in which *L* is of Type a.

**Figure 1:**
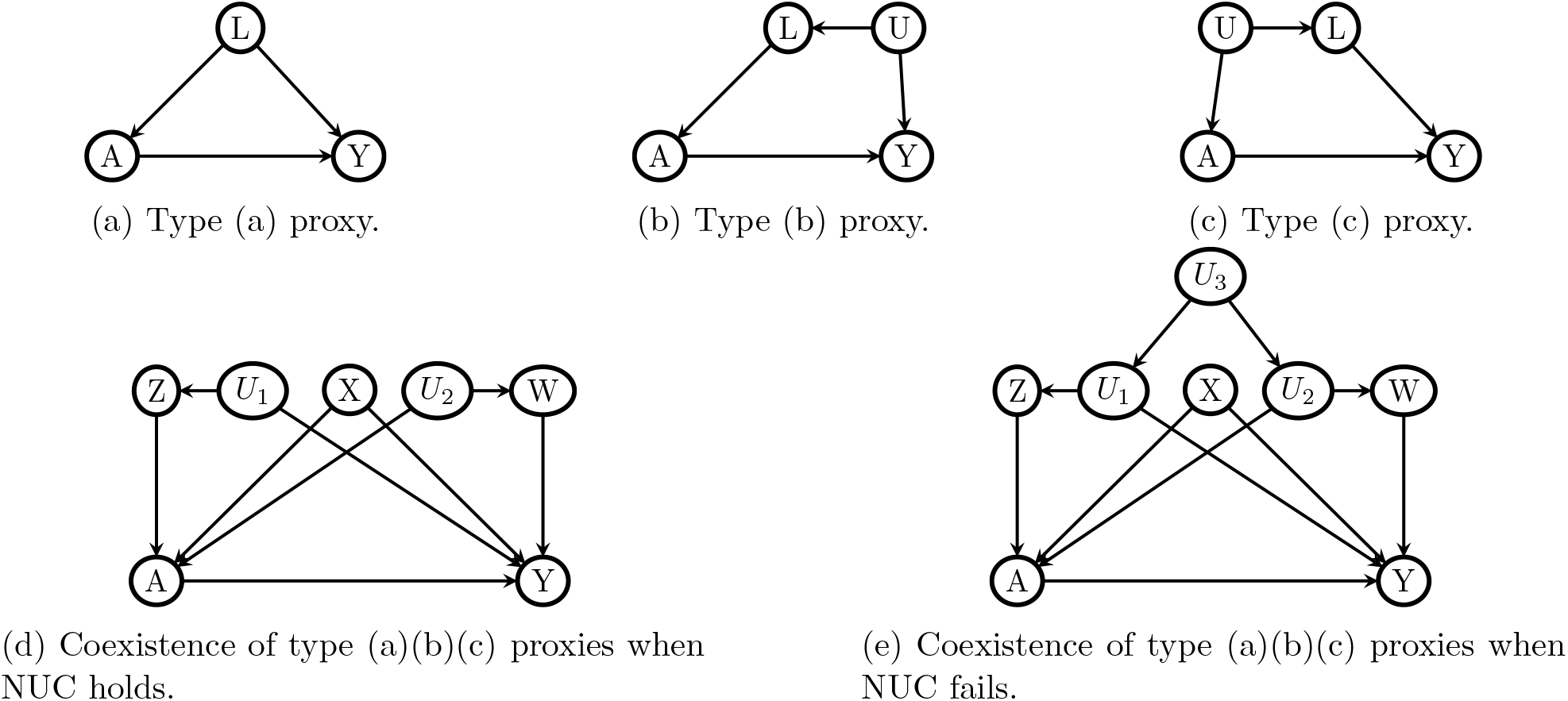
Directed Acyclic Graphs illustrating treatment and outcome confounding proxies.

Under assumptions (3)-(5), it is well-known that

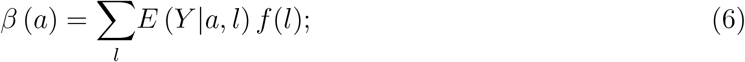

a formula most commonly known in the field of epidemiology as the g-formula^1^, a name associated with the work of James Robins which we shall adopt in this paper.

It is interesting to consider alternative data generating mechanisms under which assumption (4) holds, illustrated in Figures 1.b. and 1.c, with the first of Type b where *L* includes all causes of *A* that share an unmeasured common cause *U* (and therefore are associated) with *Y*; while the second is of Type c where *L* includes all causes of *Y* that share an unmeasured common cause *U* (and therefore are associated) with *A*. These three possibilities may coexist, as displayed in Figure 1.d. in which *L* has been decomposed into three bucket types of measured covariates *L* = (*X, W, Z*), such that *X* are measured covariates of Type a, *Z* are measured covariates of Type b, while *W* are measured covariates of Type c. At any rate, all settings represented in Figure 1 illustrate possible data generating mechanisms under which exchangeability assumption (4) holds, without necessarily requiring that the analyst identify which bucket type each covariate in *L* belongs to. Importantly, all settings given in Figure 1 rule out the presence of an unmeasured common cause of *A* and *Y*, therefore ruling out unmeasured confounding. Note that in order for exchangeability to hold in Figure 1.d, it must be that as encoded in the DAG, unmeasured variables *U*_1_ and *U*_2_ are independent conditional on *A, X, Z* and *W*; otherwise, as illustrated in Figure 1.e. the unblocked backdoor path *A* − *U*_2_ − *U*_3_ − *U*_1_ − *Y* would invalidate assumption (4). As we show in the next section, it is sometimes possible to relax this last assumption and therefore exchangeability condition (4) while preserving identification of *β* (*a*) despite the presence of unmeasured confounding, provided that one can correctly identify which bucket type each measured covariate falls into.

## 3 Proximal identification in point exposure studies

Next, consider Figure 2 which depicts a setting in which exchangeability condition (4) fails, despite having measured covariates *L* = (*X, Z, W*), owing to the presence of an unmeasured common cause *U* of *A* and *Y*. This DAG may be viewed as generalization of Figure 1.e by letting *U* = (*U*_1_, *U*_2_, *U*_3_). In the following, we propose to replace the untestable assumption (4) with an assumption that the analyst has correctly identified variables in bucket Types b and c. Formally, the causal DAG Figure 2 implies that

**Figure 2:**
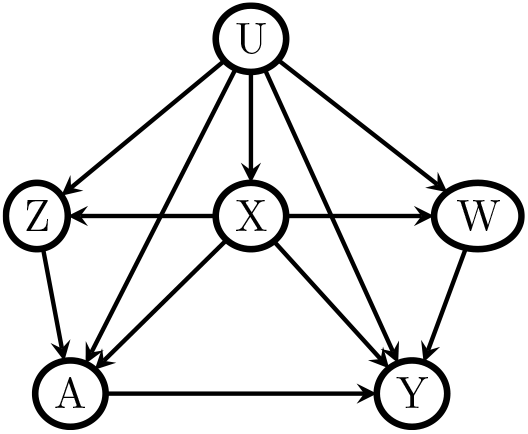
A DAG with endogenous point exposure and proxies.

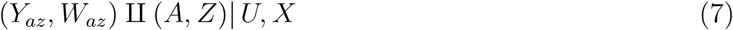

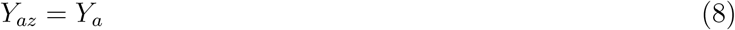

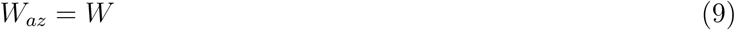

The first condition in the above display formally encodes the assumption that adjusting for (*X, U*) would in principle suffice to identify the joint causal effect of (*A, Z*) on *Y* and *W* respectively. This assumption is reasonable as long as there exist a *U* sufficiently enriched to include all common causes of (*A, Z*) and (*Y, W*) not included in *X*. As it is not required that *U* be observed, the assumption will generally hold even in observational studies. The second assumption in the above display states that *Z* does not have a direct effect on *Y* upon intervening on *A*; likewise, the third assumption states that *A* and *Z* do not have a causal effect on *W*. The first assumption will hold, provided that all variables in *Z* are correctly classified as of Type b, while the second assumption will hold provided that variables in *W* are correctly classified as of Type c. These three assumptions imply that

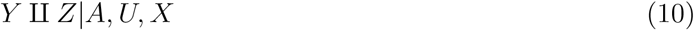

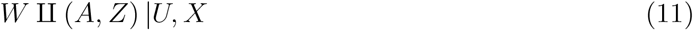

We formally refer to Type b variables *Z* as treatment-inducing confounding proxies and Type c variables *W* as outcome-inducing confounding proxies, provided that they satisfy assumptions (10) and (11). It is important to note that *Z* may not be a direct cause of *A* (in which case *Z* → *A* edge can be omitted in Figure 2), and likewise *W* may not be a direct cause of *Y* (in which case *W* → *Y* edge can be omitted in Figure 2), however as long as both are *U*-relevant, and satisfy and (11), they are considered valid proxies for our purposes and may be allocated as type b or c at the analyst’s discretion. We generally favor assumptions (10) and (11) to (7)-(9) as primitive identification conditions as they do not require conceptualizing a potential intervention on covariates *Z*.

### Remark 1.

In prior work, variables of Types b and c satisfying assumptions (10) and (11) have been called negative control exposure and negative control outcome variables, referring to negative control variables an investigator would need to supplement her observational study sample with, such that (10) and (11) would be satisfied. In this paper, we prefer the proxy terminology to the negative control nomenclature, to highlight the key observation, that often, covariates measured in an observational study in an effort to control for confounding, may not be sufficient to fulfil exchangeability, but nevertheless can potentially be partitioned into proxies satisfying negative control conditions (10) and (11). This observation, therefore alleviates the need to supplement one’s observational study design by collecting additional data on potential negative control variables, although variables of Type b and c may be enriched with appropriately selected negative control auxiliary variables when available.

### Remark 2.

It is further important to note that while we have taken Figure 2 as canonical representation of proxy variables of Types b and c, several alternative DAGs might be compatible with assumptions (10) and (11), as illustrated in the Supplemental Appendix Table A.1. Interestingly, the DAG given in first row and first column of Table A.1 of the appendix establishes that an instrumental variable (IV) for the causal effect of *A* on *Y* may be included in Type a bucket provided that it is also a valid IV for *W*. In fact, even an invalid instrumental variable which fails to satisfy the IV independence assumption^10,11^ may also be included in bucket type b as (10) and are satisfied^12^.

### Remark 3.

Additionally, one should note that similar to exchangeability condition (4), Assumptions (10) and (11) are not empirically testable as they presume certain null causal effects and involve conditional independence statements given the unmeasured variable *U*. Interestingly, the joint exclusion restriction *W*_*za*_ = *W* is a given in instances where *W* and *Z* are contemporaneous (and therefore cannot cause each other), and as assumed throughout are pre-treatment covariates. The treatment can therefore not have a causal effect on *W* as the future cannot cause the past. It is sometimes reasonable to include post-outcome variables in *Z* so that exclusion restriction conditions hold, again due to temporal ordering. However, in order to satisfy conditions (10) and (11), there must be no unblocked causal pathway between (*Y, W*) and *Z* conditional on *U, X* and *A*. Likewise, it is sometimes possible to include pre-treatment measurements of the outcome in view as potential negative control outcome and therefore to include them in *W*, provided that they satisfy conditions (10) and (11) and therefore do not have a direct effect on treatment and negative control exposure variables^12−14^. An example was provided in Miao and Tchetgen Tchetgen^12,13^ in context of studying the causal effect of air pollution on say mortality or elderly hospitalization using time series data, in which case air pollution measurement post-hospitalization may be a reasonable choice of negative control exposure to include in *Z*, and hospitalization measurement pre-air pollution may likewise be a reasonable negative control outcome to include in *W*.^12,13^

The aforementioned connection to negative control literature is instrumental in determining sufficient conditions for nonparametric identification of *β* (*a*) by leveraging identification results recently obtained by Miao et al.^15^ We summarize their results below, provide intuition for the results and refer the interested reader to their manuscript for a careful treatment of mathematical conditions underpinning the approach. In the next Section, we extend their results to the time-varying setting, which to our knowledge is new to the literature, all proofs can be found in the Appendix.

Let *h* (*a, x, w*) denote a solution to the equation:

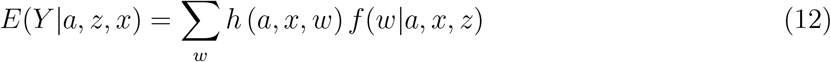

where in slight abuse of notation ∑ denotes an integral in case of continuous *w*. Next, suppose that the following conditions hold, for any function *υ* (·):

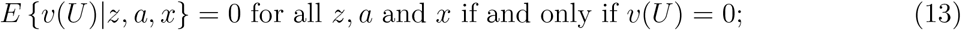

This condition is formally referred to as a completeness condition which accommodates both categorical and continuous confounders. Completeness is a technical condition taught in most foundational courses in theory of statistical inference. Here one may interpret it as a requirement relating the range of *U* to that of *Z* which essentially states that the set of proxies must have sufficient variability relative to variability of *U*. The condition is easiest understood in the case of categorical *U, Z* and *W*, with number of categories *d*_*u*_, *d*_*z*_ and *d*_*w*_ respectively. In this case, completeness requires that

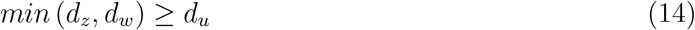

which states that *Z* and *W* must each have at least as many categories as *U*. Intuitively, condition (14) states that proximal causal learning can potentially account for unmeasured confounding in the categorical case as long as the number of categories of *U* is no larger than that of either proxies *Z* and *W* ^16^. This further provides a rationale for measuring a rich set of baseline characteristics in observational studies as a potential strategy for mitigating unmeasured confounding via the proximal approach we now describe. Miao et al^15^ established that under assumptions (10)-(13), the counterfactual mean *β* (*a*) can be identified nonparametrically by the formula:

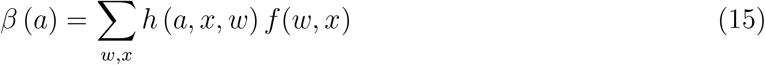

We refer to equation (15) as the *proximal g-formula*, and to *h* (*a, x, w*) as an outcome confounding bridge function^15^. A few key observations are in order. First, equation (12) defines a so-called inverse problem formally known as a Fredholm integral equation of the first kind. Formal conditions for existence of a solution of such an equation are well established in functional analysis in mathematical literature, but due to their technical nature are beyond the scope of the current paper, though it is worth mentioning that existence of a solution requires the following additional completeness condition:

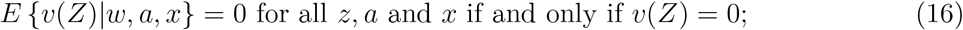

An assumption that cannot hold unless *W* is *U*-relevant. We refer the reader to Miao et al for a technical exposition of required regularity conditions. In the categorical case, as established in Shi et al^16^ condition (14) along with a rank condition for a certain matrix defined in terms of the conditional distribution of *W* given (*Z, A, X*) suffices for equation (12) to admit a solution^16^. It is important to note that *h* (*a, x, w*) satisfying (12) need not be unique, any solution to this equation yields the same value of the proximal g-formula. We also note that by latent exchangeability, *β* (*a*) = ∑_*u,x*_ *E*∑(*Y* |*a, u, x*) *f* (*u, x*) = ∑_*w,x*_ *h* (*a, x, w*) *f* (*w, x*); and as shown in by Miao et al^15^, ∑*E* (*Y*_*a*_|*u, x*) = ∑ _*w*_ *h* (*a, x, w*) *f* (*w*|*u, x*), which highlights the inverse-problem nature of the task accomplished by proximal g-formula, which is to determine an *h* that satisfies this equality without explicitly modeling or estimating the latent factor *U*. A remarkable feature of proximal learning is that accounting for *U* without either measuring *U* directly or estimating its distribution can be accomplished provided that the set of proxies though imperfect, is sufficiently rich so that the inverse-problem admits a solution in a model-free framework.

Intuition about conditions under which a unique solution to equation (12) might exist can be gained in the simple case of binary *A, W, Z*, whereby it is straightforward to show that the unique solution to (12) is

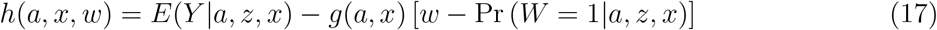

Where

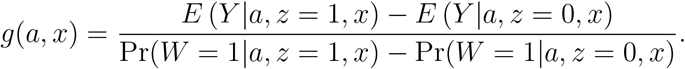

Importantly, note that although the right-hand side to equation (17) appears to depend on *z*, the left-hand side indicates that it does not, which is readily verified with some algebra. In order for *h* to be finite, one requires that Pr(*W* = 1 | *a, z* = 1, *x*) − Pr(*W* = 1 | *a, z* = 0, *x*) ≠ 0; that is *W* must be associated with *Z* conditional on (*A, X*), a condition that one would expect to hold to the extent that *W* and *Z* are strong proxies of *U*, thus further highlighting the importance of selecting strong potential proxies. In the binary case, *β* (*a*) takes the closed form:

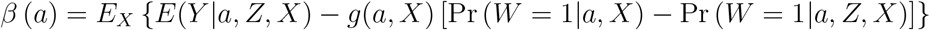

A generalization of the above closed-form expression for proximal g-formula with categorical variables is given in Shi et al^16^ for the average causal effect *β* (1) − *β* (0) of a binary treatment on the additive scale. Unfortunately, unlike the g-formula, the proximal g-formula is not always available in closed-form and requires solving equation (12) numerically, which might be computationally intensive and unstable due to its potential to be empirically ill-posed. Ill-posedness in this case refers to the fact that small amount of uncertainty in estimating the left handside of (12) empirically can often induce excessive uncertainty in obtaining a solution to the equation. Such ill-posedness is typically addressed by some form of regularization of the integral equation. Below, we describe a simple statistical modeling approach analogous to g-computation, which sidesteps this difficulty by automatically generating stable solutions to the equation under correct model specification. Revisiting the motivating example given in the introduction, one may readily verify that the structural equations (1) imply that there exists coefficient 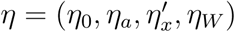 such that:

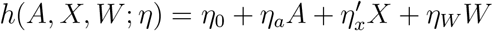

and

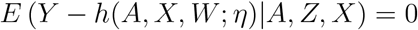

so that equation (12) is satisfied. By then applying the proximal g-formula, one recovers *β*_*a*_ = *η*_*a*_ = *E* {*h*(*A* = 1, *X, W*; *η*) − *h*(*A* = 0, *X, W*; *η*)} identifies the causal effect parameter.

It is interesting to compare proximal g-formula to standard g-formula^1^. In this vein, suppose that *L* = (*X, W*) suffices for exchangeability condition (4), so that *Z* = Ø; then, proximal g-formula reduces to the standard g-formula with *h* (*a, x, w*) = *E*(*Y* | *a, x, w*) a stable solution to (12) given that:

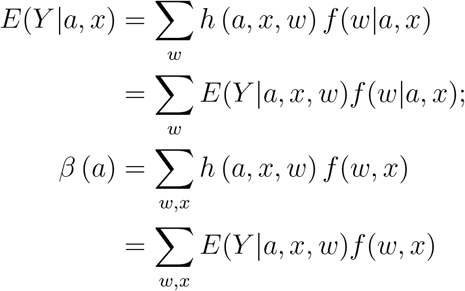

From this perspective, exchangeability may be viewed as a form of regularization of equation (12) which automatically yields a unique stable solution to the integral equation.

### Remark 4.

We note that Miao and Tchetgen Tchetgen^12^ considered alternative identifying conditions in that instead of taking equation (12) as starting point, they a priori assume that there exist a bridge function *h*(*w, a, x*) such that *E*(*Y*_*a*_ | *u, x*) = ∑_*w*_ *h* (*a, x, w*) *f* (*w* | *u, x*); in addition, they replace completeness condition (13) which is not subject to an empirical test, with the testable completeness condition that {*E v*(*w*) | *z, a, x*} = 0 for all *z, a* and *x* if and only if *v*(*w*) = 0; then they establish that such function *h* must solve equation (12).

## 4 Proximal identification in complex longitudinal studies

We now consider proximal identification of causal effects in complex longitudinal studies. In order to ground ideas and simplify the exposition, we focus primarily on a special case of a longitudinal study with two follow-up times and briefly review identification under a longitudinal version of exchangeability. Thus, suppose that one has observed time-varying treatment and covariate data {*L* (*j*), *A* (*j*)} at follow up visits *j* = 0, 1 of a longitudinal study. Let *Y* denote the outcome of interest measured at the end of follow-up *j* = 2. We assume that recorded data on the treatment and prognostic factors do not change except at these times, moreover, *L*(*j*) temporally precedes *A*(*j*). We use overbars to denote the history of that variable up to end of follow-up; for example 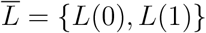. Let *Y*_*ā*_ = *Y*_*a*(0),*a*(1)_ denote the potential outcome had possibly contrary to fact, a subject followed treatment regime *Ā* = *ā*. Our aim is to identify the potential outcome mean *β* (*ā*) = *E* (*Y*_*ā*_). To do so, three standard assumptions are typically invoked. The first entails a longitudinal version of consistency:

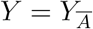

linking counterfactual outcomes {*Y*_*ā*_ : *ā*} to observed variables (*Y, Ā*). The next assumption is that there are no unmeasured confounders for the effect of *A*(*j*) on *Y*, that is, for all treatment histories *ā*,

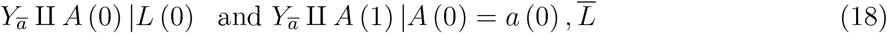

This assumption which generalizes exchangeability to the longitudinal setting, is also known as the sequential randomization assumption (SRA)^17^. It states that conditional on treatment history and the history of all recorded covariates up to *j*, treatment at *j* is essentially randomized by nature and thus must be independent of the counterfactual random variable *Y*_*ā*_.^2,17^

We finally assume that the following positivity assumption holds. For all *a*(*j*) in the support of *A*(*j*)

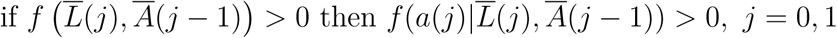

with *A*(−1) ≡ 0, which essentially states that if any set of subjects at time *j* have the opportunity of continuing on a treatment regime *a* under consideration, at least some will take that opportunity. Robins established that under these assumptions, the counterfactual mean *β* (*a*) is given by the longitudinal g-formula^2,17^:

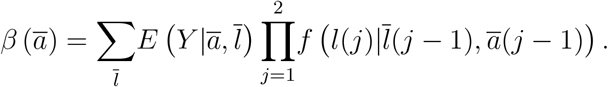

As argued in the introduction, in an observational study, the assumption of no unmeasured confounding cannot be guaranteed to hold, and it is not subject to empirical test, even when good efforts are made to collect data on crucial covariates. As before, our aim is to relax sequential exchangeability/SRA by explicitly incorporating measured covariates as proxies of underlying confounding mechanisms longitudinally. In this vein, we suppose that one can partition covariates measured at time *j* = 0, 1 into three bucket types *L*(*j*) = (*Z*(*j*), *W* (*j*), *X*(*j*)), *j* = 0, 1, such that the following conditions hold:

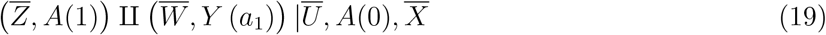

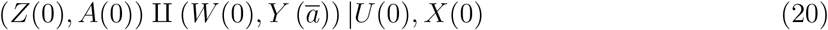

These conditions are a longitudinal generalization of (7).

Figure 3 illustrates a possible data generating mechanism in which conditions (11) and (20) hold, where to simplify the figure certain edges are shaded and we have suppressed observed time-varying covariates 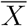 which structurally follow the same relationship as *Ū* with other variables. Additionally, for identification we require the following longitudinal generalization of completeness condition (13), which state that for any function *υ*(·):

**Figure 3:**
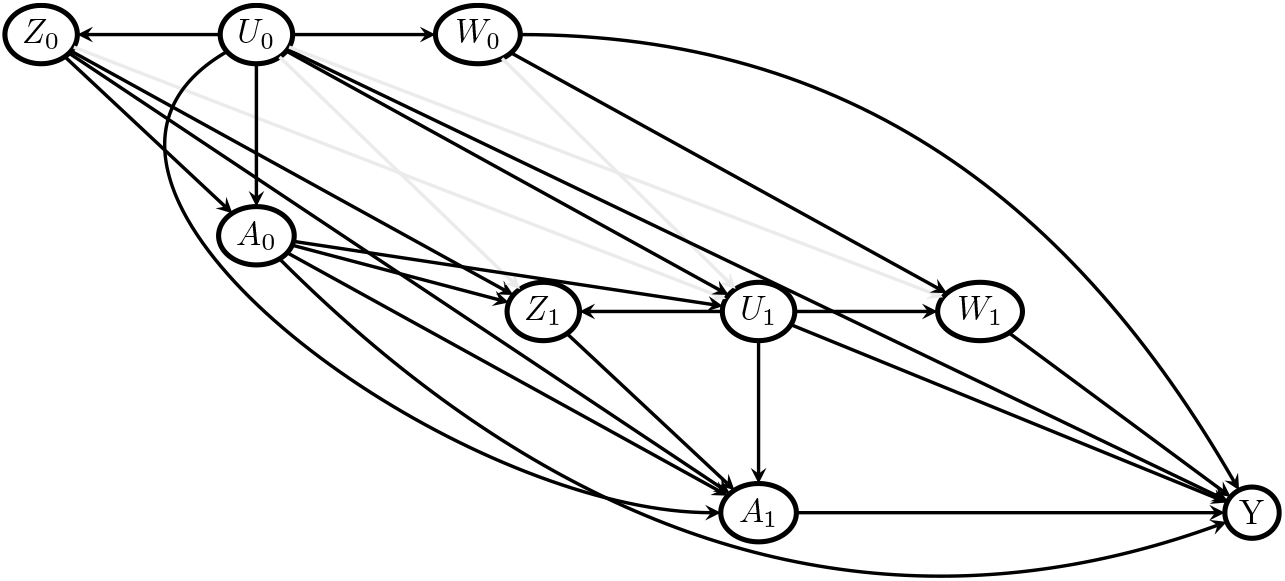
A DAG with endogenous time varying treatments and proxies.

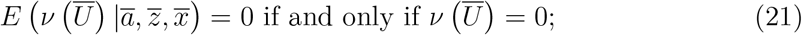

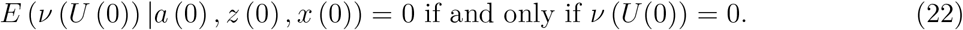

Finally, extending assumption (12) to longitudinal setting, we suppose that there exist functions 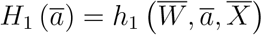 and *H*_0_ (*ā*) = *h*_0_ (*W* (0), *ā, X* (0)) that solve equations:

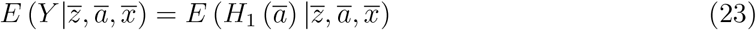

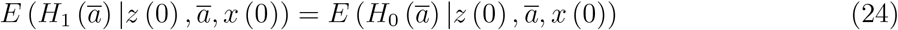

Confounding bridge equation (23) is exactly equivalent to equation (12) with 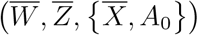 replacing (*W, Z, X*), and *A*(1) replacing *A*. As show in the result below, this assumption yields identification of 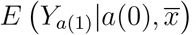, while the second assumption (24) which does not have a point exposure analog, yields identification of *E* (*Y ā*| *x*(0)). In fact, we have the following result.

### Result 1

Suppose that assumptions (19)-(24) are satisfied, then we have that

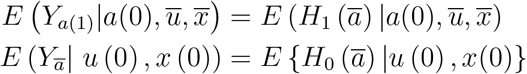

and

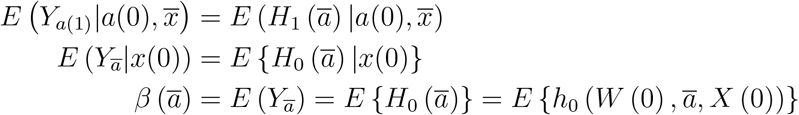

Result 1 can be extended to a longitudinal study of follow-up length *J* > 2 as shown in the Appendix; the proof of which implies Result 1 as a special case. As in the point treatment setting, *H*_1_ (*ā*) and *H*_0_ (*ā*) need not be uniquely identified in order for *β* (*ā*) to be uniquely identified.

## 5 Proximal g-computation

In this section we describe a practical approach for estimating the proximal g-formula. we first describe the approach in the point treatment case before extending it to the case of time-varying treatment. Thus, suppose that one has observed an i.i.d sample of size *n* on (*A, L* = (*X, Z, W*)). It is then convenient to directly specify a parametric model for the outcome bridge function:

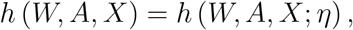

with unknown parameter *η*; and for the joint law

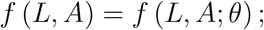

with unknown parameter *θ*. Note that together, these models entail a parametric model for

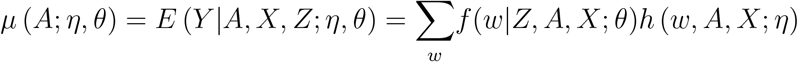

in terms of *θ* and *η*. This modeling assumption is therefore appropriate only if the outcome mean admits the representation given above. As we show below, directly modeling the outcome confounding bridge function obviates the need to solve complicated integral equations which are well-known to be ill-posed and therefore to admit unstable solutions. The above modeling strategy can be viewed as a form of regularization of the problem so as to resolve ill-posedness. Although not pursued here, a variety of semiparametric (e.g. partially linear model, single index model) or nonparametric (e.g. generalized additive, reproducing kernels, neural networks) may be used to model *h* more flexibly, thus alleviating concerns about specification bias.

Let 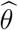 denote the maximum likelihood estimator of *θ*, and define 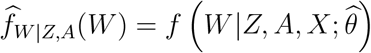 implied by 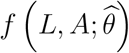 One may then estimate *η* based on Result 1, by fitting via least-squares, the regression model: *µ* (*A, η*) = *E* (*Y* |*A, X, Z*) = *E* (*H* (*η*) |*A, X, Z*) given by

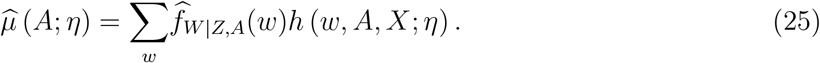

For continuous *Y*, this may be accomplished by least-squares minimization:

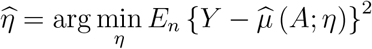

where *E*_*n*_ stands for sample average. Then, assuming all models are correctly specified, one can show that 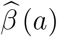 is a consistent and asymptotically normal estimator of *β* (*a*), where

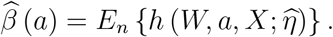

For inference, we recommend using the nonparametric bootstrap to obtain standard errors and confidence intervals. We note that evaluating (25) might require evaluating either a sum, an integral or both with respect to a high dimensional variable *w*; in many cases, the sum/integral may not admit a closed form expression or may be computationally prohibitive to evaluate, in which case Monte Carlo approximation of 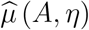 may provide a practical solution. In case of binary *Y*, use of a link function (say logit or probit link function) may be necessary in specifying model for *h* (*w, A, X*; *η*), in order to ensure that *µ* (*A*; *η, θ*) = Pr (*Y* = 1 | *A, Z, X*; *η, θ*) lies in the unit interval (0,1). Estimation in the binary case can then proceed by standard maximum likelihood estimation thus maximizing the log likelihood function

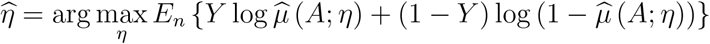

In the Appendix, we describe special cases where 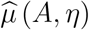 admits a closed form expression. Here, we discuss the important special case of proximal g-computation under a linear specification for *h* (*W, A, X*; *η*), say

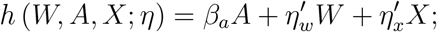

so that *E*(*Y*_*a*_) = *β* (*a*) = *β*_0_ + *β*_*a*_*a* where 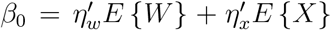 where 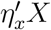 in includes an intercept term. Suppose further that one specifies a (multivariate) linear regression model

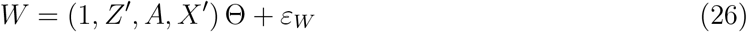

Then, one can estimate the average causal effect *β*_*a*_ = *E*(*Y*_*a*+1_ − *Y*_*a*_) with the regression coefficient 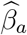 obtained by fitting the standard linear regression model:

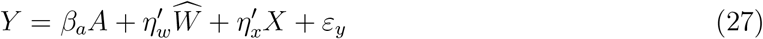

by least-squares, where 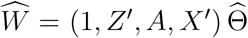 is the element-wise least-squares regression of *W* on (1, *Z* ′, *A, X*′)^12^. We refer to this procedure as proximal two-stage least squares (P2SLS) given its close relationship to 2SLS estimation in instrumental variable setting^18^. This connection in fact has implications for practice as it indicates that the estimator can be implemented with any off-the-shelf instrumental variable software which can perform 2SLS for multivariate exposure variable upon taking *W* as the endogenous (multivariate) variable, *Z* playing the role of IV, with *A* taken as a covariate. Such software can be used to obtain 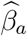 and corresponding confidence intervals, accounting for the uncertainty in the first stage estimator 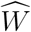. The model given in equation (27) has an interesting interpretation as it emulates a standard regression adjustment of confounding by *X*, and further adjusts for 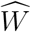 as proxy for the unmeasured factor *U* therefore deconfounding the standard regression approach. As mentioned in the introduction, we can therefore refer to 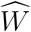as *proximal control variable*.

### Remark 5.

One may note from the description of P2SLS that in the event that dim(*Z*)<dim(*W*), *η*_*w*_ in (27) may not be uniquely identified; nonetheless, it is straightforward to verify that all least squares solutions for 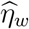 yield a consistent estimator 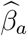 Either way, a test of presence of confounding bias entails a standard statistical test of the null hypothesis that all components of *η*_*w*_ are identically zero, which is readily available even when dim(*Z*)<dim(*W*).

Next, we consider the longitudinal setting where we observe an i.i.d sample of size *n* on 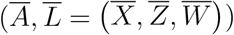. Then, parametric proximal g-computation relies on specifying parametric models for the outcome bridge functions:

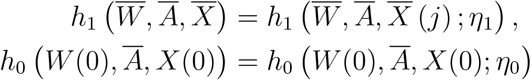

with unknown parameter *η*_*j*_; and for the joint law

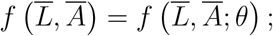

with unknown parameter *θ*. Let 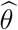 denote the maximum likelihood estimator of *θ*, and define 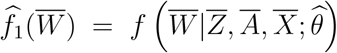 and 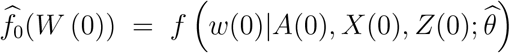 both deduced from 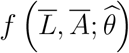. Then we propose to estimate *η*_*j*_ based on Result 1, by recursively fitting regression models of *Y* on 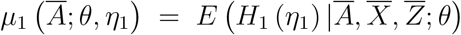, and of *H*_1_ (*η*_1_) on *µ*_0_ (*Ā*; *η*_0_, *θ* = *E* (*H*_0_ (*a*_1_, *η*_0_) |*A*(0), *X*(0), *Z*(0); *θ*) given by

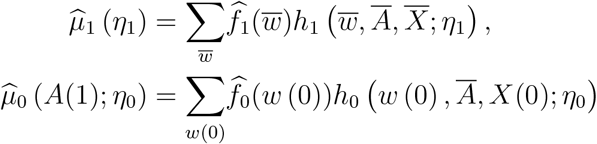

Each of these regressions can readily be performed via ordinary least-squares. Then, assuming all models are correctly specified and Result 1 holds, one can show that 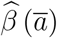 is a consistent estimator of *β* (*ā*), and is approximately normally distributed, where

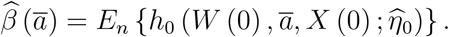

In order to estimate standard errors for 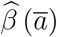 and confidence intervals for *β* (*ā*), we recommend using the nonparametric bootstrap^19^. We refer to the above estimation procedure as parametric proximal g-computation, the proximal analog to parametric g-computation algorithm of Robins^2,17^.

The algorithm simplifies tremendously in case of additive confounding bridge functions, say:

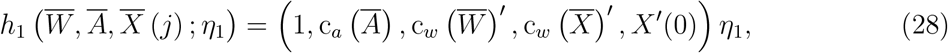

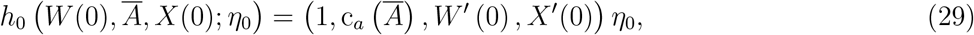

where for time-varying variable 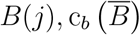 denotes a user specified function 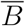 of for instance, we might take 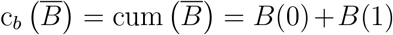, where in case of vector *B*, the sum applies entry-wise such that 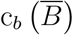 is a vector of the same dimension as *B*(*j*). Then proximal g-computation can be implemented by the following recursive least-squares algorithm:

### Proximal recursive least squares algorithm

Step 1: fit the multivariate linear regression

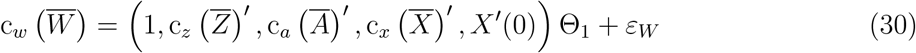

by applying least-squares separately to each entry of vector 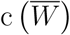, and let

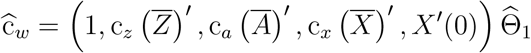

denote its fitted values;

Step 2: fit the linear regression

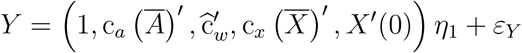

by least-squares where we note that 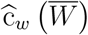 has been substituted in for 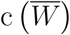, and let

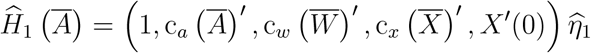

Step 3: fit the multivariate linear regression

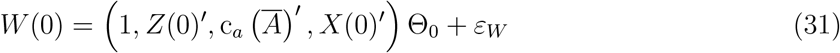

by applying least-squares separately to each entry of vector *W* (0), and let

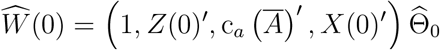

denote its fitted values; fit the linear regression

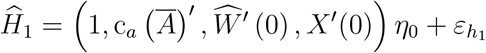

by least-squares, to obtain an estimate of *H*_0_,

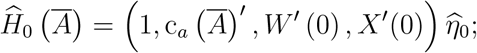

Step 4:Evaluate

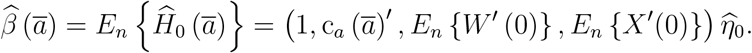

It is important to note that although additive, the specific form of models used in Steps 1-4 can be quite flexible and can accommodate both nonlinearities (e.g. using either polynomial or splines to model covariates) as well as interactions with treatment or among covariates. More flexible models can be somewhat more involved as each nonlinear specification of 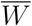 entries requires a corresponding regression in Step 1. We also note that the particular manner in which a treatment or covariate history enters a given model is entirely to the discretion of the analyst. For instance, natural options for c_*a*_ *Ā* include (*A*(0), *A*(1), *A*(0) × *A*(1)), cum (*Ā*) or simply *A*(1) as viable alternatives depending on their respective goodness-of-fit.

Interestingly, under linearity of *H*_0_ and *H*_1_ with respect to 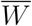, the proximal recursive least squares algorithm yields an estimator of *β* (*a*), that remains consistent even if linear models (30) and (31) for 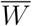 and *W* (0) are misspecified. Likewise, in the point treatment setting, 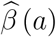 can be shown to remain consistent even if model (26) is not correctly specified provided that *h*_0_ and *h*_1_ are correctly specified. The implication of this result is that OLS provides extra protection against model misspecification bias in modeling 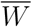 as a linear model, including for binary or discrete components of *W*. This is an important property that does not generally hold for proximal g-computation algorithm which requires in addition to correct specification of *h*_0_ and *h*_1_, that one also specify 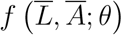 correctly. It is however possible to obtain an estimator of *h*_0_ (*η*_0_) and *h*_1_ (*η*_1_) using a recursive generalized methods of moments (RGMM) which does not require a model for 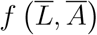 and therefore is not susceptible to bias due to modeling the latter incorrectly. We refer the interested reader to Miao and Tchetgen Tchetgen^12^ in point treatment case. A detailed treatment of this more robust estimation approach in longitudinal settings will be described elsewhere. It is worth noting that when 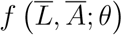 is correctly specified, one can generally expect proximal g-computation to be more efficient than proximal recursive least square and recursive generalized method of moments.

## 6 Data applications

### 6.1 Point treatment application

We first illustrate proximal estimation of causal effects in a point treatment application to the Study to Understand Prognoses and Preferences for Outcomes and Risks of Treatments (SUPPORT) with the aim of evaluating the causal effect of right heart catheterization (RHC) during the initial care of critically ill patients in the intensive care unit (ICU) on survival time up to 30 days^20^. RHC was performed in 2184 patients within the initial 24 hours of ICU stay, while 3551 patients were managed without RHC. The SUPPORT study collected rich patient information encoded in 73 covariates, including demographics (such as age, sex, race, education, income, and insurance status), estimated probability of survival, comorbidity, vital signs, physiological status, and functional status. The outcome of interest is the number of days between admission and death or censoring at 30 days (*Y*). Ten variables measuring the patient’s overall physiological status were measured from a blood test during the initial 24 hours in the ICU: `serum sodium, serum potassium, serum creatinine, bilirubin, albumin, PaO2/(.01*FiO2) ratio, PaCO2, serum PH (arterial), white blood cell count`, and `hematocrit`. These variables may be subject to substantial measurement error and as single snapshot of underlying physiological state over time may be viewed as potential confounding proxies. Among the ten physiological status measures, four `(pafi1, paco21, ph1, hema1)` are strongly correlated with both the treatment and the outcome; thus we construct proxies *Z* and *W* from this reduced set of variables and collect all 67 remaining variables as covariates (*X*).

As we hypothesize that our four candidate proxies are equally likely to be valid treatment-inducing proxy or outcome-inducing proxy, we consider a practical strategy for allocating the four candidate proxies to bucket types b and c. The approach first ranks proxies according to their strength of association in treatment (based on logistic regression of *A* on *L*) and outcome models (based on linear regression of *Y* given *A* and *L*) respectively; next, we select proxies in decreasing order of strength of association, first selecting the proxy with strongest association with the outcome as outcome-inducing proxy and likewise for the treatment. In case of a tie, that is if these are the same variable, one may either decide to prioritize one of the two buckets or alternatively to randomize allocation to a proxy bucket type; upon allocating a given variable, say to the outcome-inducing proxy bucket, one subsequently removes the variable from the list of remaining treatment-inducing candidate proxies, and vice-versa. The algorithm stops when all proxies have been allocated. The algorithm produced the allocation *Z* = (`pafi1, paco21`) and *W* = (`ph1, hema1`). For estimation of the causal effect of interest, we assume a linear outcome confounding bridge function,

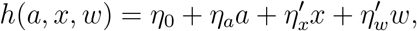

in which case, the coefficient *η*_*a*_ = *β*_*a*_ = *E* {*h*(1, *X, W*) −*h*(0, *X, W*)} encodes the causal effect of interest. After allocating the proxies, we apply two stage least squares to estimate the confounding bridge function, which can be implemented via routine R software such as `ivreg`. The following is a standard call of `ivreg` in R,

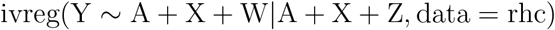

Ordinary least squares results in a negative and statistically significant causal effect estimate 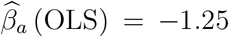 with standard error=0.28. Outcome-inducing proxy ph1 is associated with confounding bridge parameter (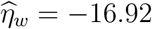, standard error=8.8), indicating moderate empirical evidence that unmeasured conf ounding might be biasing 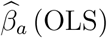 (OLS). The causal effect estimate obtained by P2SLS is substantially larger than standard OLS point estimate 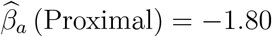 with corresponding standard error= 0.43. These results suggest that RHC may have an even more harmful effect on 30 day-survival among critically ill patients admitted into an ICU than previously documented. Results from this analysis are summarized in tables provided in the Supplemental Appendix.

### 6.2 Time-varying treatment application

We reanalyze data from an article published by Choi et al (2002)^21^ on the potential protective effects of the anti-rheumatic therapy Methotrexate (MTX) among patients with rheumatoid arthritis. While Choi et al focused on survival as an endpoint and used a Cox marginal structural models to quantify joint treatment effects under SRA, here we consider the joint causal effects of MTX on average of reported number of tender joints, an important measure of disease progression, without appealing to SRA. Our analysis includes individuals who were older than age 18 years and who attended the Wichita Arthritis Center at least twice between Jan 1, 1981 (when weekly low-dose methotrexate therapy and health assessment questionnaire scores became available) and Dec 31, 1999; had rheumatoid arthritis fulfilling the 1958-1987 American College of Rheumatology (formerly the American Rheumatism Association) criteria for rheumatoid arthritis; and had not received methotrexate before their first visit to the center, who survived more than 12 months.

Methotrexate use and dose was recorded in the computer database at each clinic visit. We classified methotrexate exposure status as ever-treated or never-treated, i.e., once a patient starts methotrexate therapy, he or she was considered on therapy for the rest of the follow-up. This approach provides a conservative estimate of methotrexate efficacy just as intent-to-treat analysis does in randomized clinical trial.

A thousand and ten patients with rheumatoid arthritis met our inclusion criteria, 183 of them were treated with methotrexate at month 6 of follow-up. We have recorded baseline covariates including age, sex, past smoking status, education level, rheumatoid arthritis duration, calendar year and rheumatoid factor positive. Time varying covariates include current smoking status, health assessment questionnaire, number of tender joints, patient’s global assessment, erythrocyte sedimentation rate, number of disease modifying antirheumatic drugs taken and prednisone use. Our objective is therefore to evaluate the joint effects of MTX use at baseline and month six on average of tender joints at month 12 of follow-up. In addition to proximal learning, for comparison, similar to Choi et al, we also evaluated the causal effect of interest under a marginal structural linear model *E*(*Y*_*ā*_) = *β*_0_ + *β*_*a*_cum (*ā*) = *β*_0_ + *β*_*a*_ {*a* (0) + *a*} where *a*(0) and *a*(1) are MTX use at baseline and at month 6 respectively, estimated via standard inverse probability weighted least squares assuming SRA given both all baseline and time-varying covariates.

We then implemented proximal recursive least squares algorithm under linear outcome confounding bridge specification (28) and (29), with *X* =(age, education, sex, smoking, rheumatoid arthritis duration, calendar year). Since number of tender joints at one year of follow-up is the primary outcome, tender joints count (jc) at baseline and at follow-up month 6 are both natural candidate as outcome-inducing proxies. Other candidate proxies included health assessment questionnaire (haqc), patient’s global assessment of disease status (gsc) and erythrocyte sedimentation rate (esrc), number of disease-modifying antirheumatic drugs (dmrd), rheumatoid factor positive (rapos) and prednisone use (onprd2). We further reduced the set of candidate proxies to candidate variables associated with both treatment and outcome variables. Finally, we applied the allocation algorithm described in the prior section resulting in *Z* (*j*) = haqc (*j*) and *W* (*j*) = jc(j).

IPW least squares suggests a protective effect of MTX with 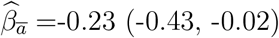, although validity of this finding is contingent on SRA. Proximal recursive least-squares yields results suggests a stronger protective effect 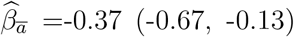, with strong evidence of confounding bias 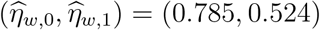 with corresponding 95% confidence intervals (0.50, 1.1) and (0.33, 0.71) respectively. These results reinforce understanding of potential protective effects of MTX on disease progression. Results from this analysis are summarized in tables provided in the Supplemental Appendix.

## 7 Discussion

We have described a new framework for the analysis of observational data subject to potential confounding bias. The approach acknowledges that in practice, measured covariates generally fail in observational settings to capture all potential confounding mechanisms and at most may be seen as proxy measurements of underlying confounding factors. Our proximal causal learning framework provides a formal potential outcome framework under which one can articulate conditions to identify causal effects from proxies. We have described proximal g-formula and proximal g-computation algorithm for estimation in point treatment and time-varying treatment settings. The proximal approach is closely related to negative control methods recently proposed for detection and sometimes estimation of point treatment interventions^12,13,16,22^. We refer the reader to Shi et al^23^ for a recent review of negative control literature.

While similar to standard g-computation, our proximal g-computation algorithm (as well as proximal two stage least squares and recursive least squares) rely on correct specification of outcome confounding bridge functions, we are currently in the process of developing alternative methods which similar to inverse-probability weighting, rely on a model for a so-called treatment confounding bridge function such that it is possible to construct two separate estimators of the average treatment effect each depending on a different model; either outcome or treatment confounding bridge function. Interestingly, we have also developed doubly robust estimators that, similar to standard doubly robust estimators developed by Robins and colleagues^24^, remain unbiased in large samples provided at least one confounding bridge function model is correct, but not necessarily both. These results along with further evaluation of finite sample performance of proximal inference will be presented in future papers.

## Data Availability

Data are available upon request to the first author

## Appendix

### Proof of Result 1

We establish the following general result which implies Result 1:suppose that for *j* = *J* − 1, …, 0

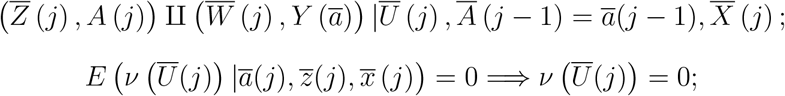

and that there exist a function 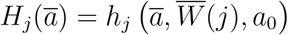 such that

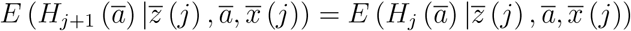

generalizations of conditions (19)-(24); then we have that

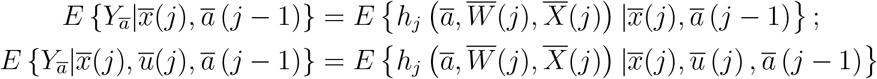

and

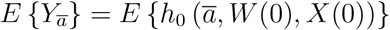

To prove the result, consider *j* = *J* − 1, then we have that

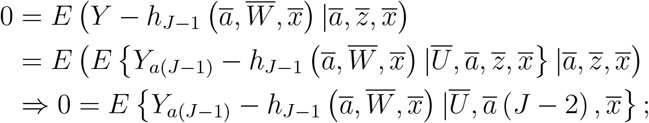

therefore

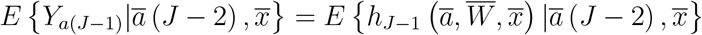

Next,

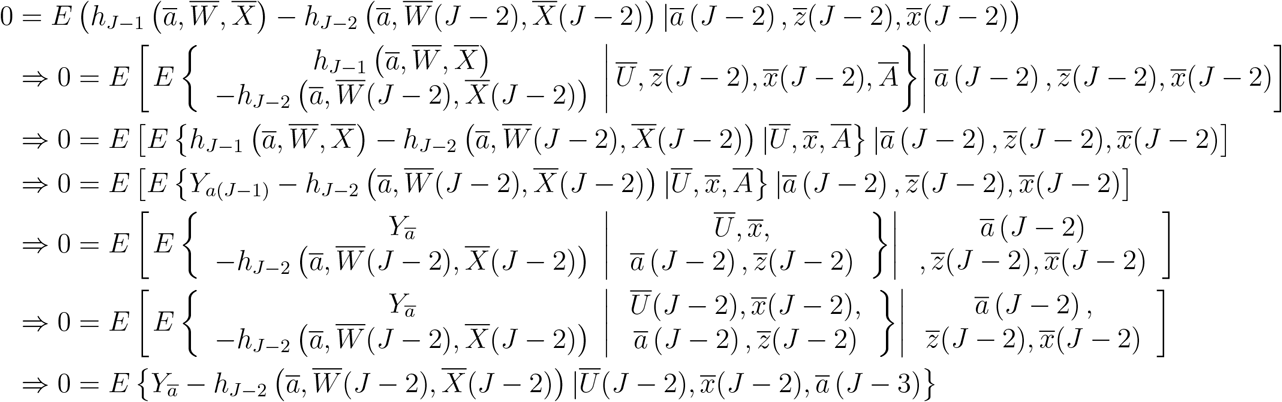

Therefore

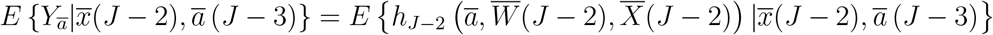

Repeating this argument for *j* = *J* − 3, 0, we arrive at

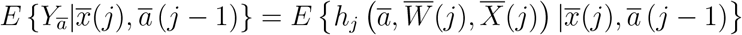

and

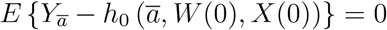

proving the result.

### Closed form expression of *µ* (*A, η*) for binary *Y*

Suppose that *Y* is binary and *W* is a continuous scalar variable. Further suppose that

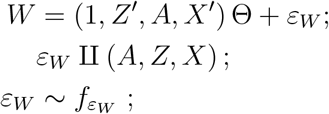

next, suppose that

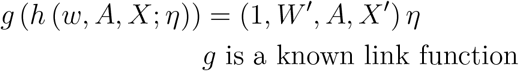

then we have that

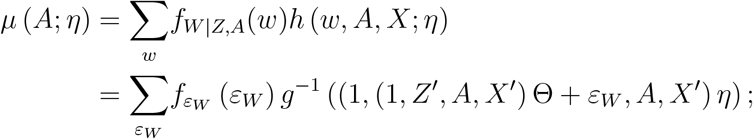

Suppose finally that one specifies 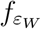 to match the bridge distribution function of the link function *g*^20^, then one can show that

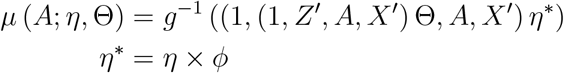

with 0 < *ϕ* < 1^20^. The form of *ϕ* depends on the bridge distribution function for the link *g*. For instance, for *g* the probit link, we have that 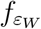 is a zero mean Gaussian density with variance *σ*^2^ and 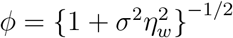 and

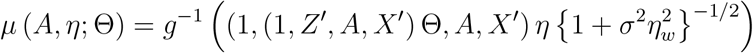

All parameters can be estimated by maximizing the log-pseudo-likelihood function

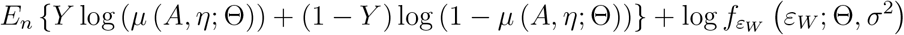

In case *g* is logit link, the above log likelihood is modified by setting 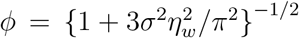 and 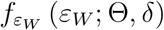 the logistic bridge function *B*_*l*_ (0, *δ*) of Wang and Louis^25^. In case *W* is multi-variate with both continuous and discrete components we factorize 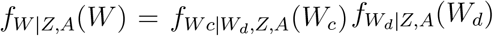 where 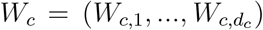 are continuous components of *W* and *W*_*d*_ are discrete components. It is then convenient to take *g* as probit link function and *W*_*c*_ *W*_*d*_, *Z, A, X* as multi-variate Gaussian with mean 0 and variance-covariance matrix Σ, in which case

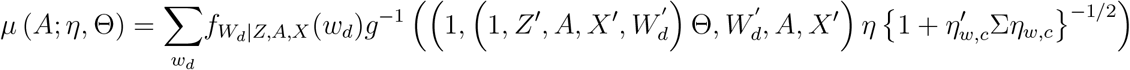

### Generalization of Proximal recursive least squares algorithm

Step 1: For user-specified functions 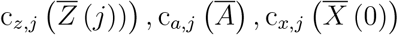 fit the multivariate linear regression

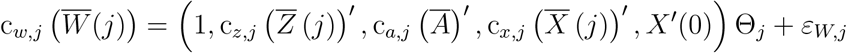

*j* = *J* − 1, …, 0 by applying least-squares separately to each entry of vector 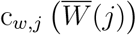, and let

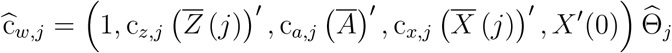

denote its fitted values;

Step 2:Let 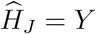 and for *j* = *J* − 1, …, 0, fit the linear regression of

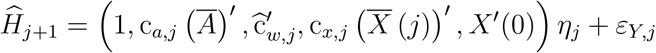

by least-squares where we note that 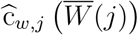has been substituted in for 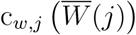, and let

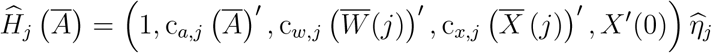

Step 3:Evaluate

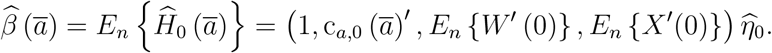

Next we show that 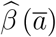 is consistent for *β* (*ā*) provided that

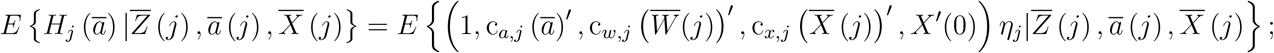

even if

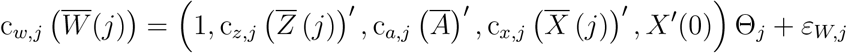

is misspecified. To prove this result, it suffices to note that

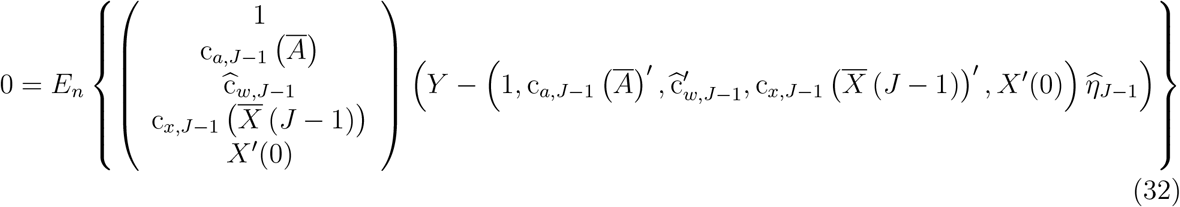

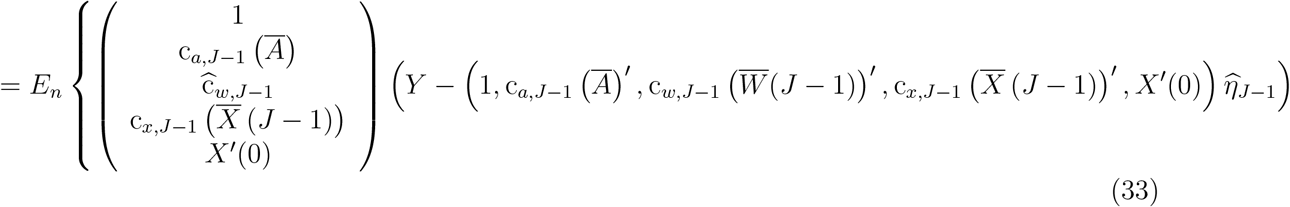

because

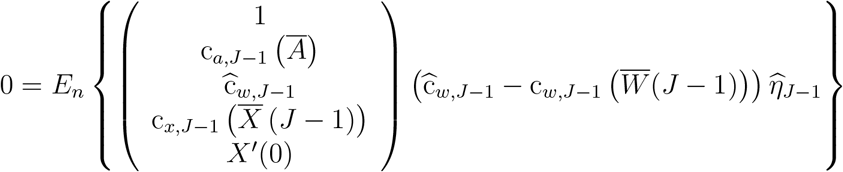

by virtue of 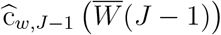 being the least-square projection of 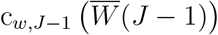 onto

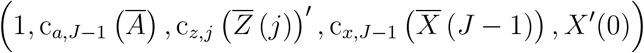

which spans

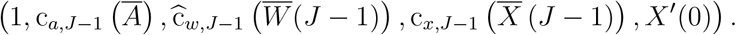

Equation (33) yields a consistent estimator of *η*_*J*−1_ because 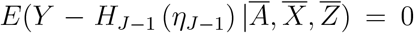. Likewise, for any *j* < *J* − 1 we have that

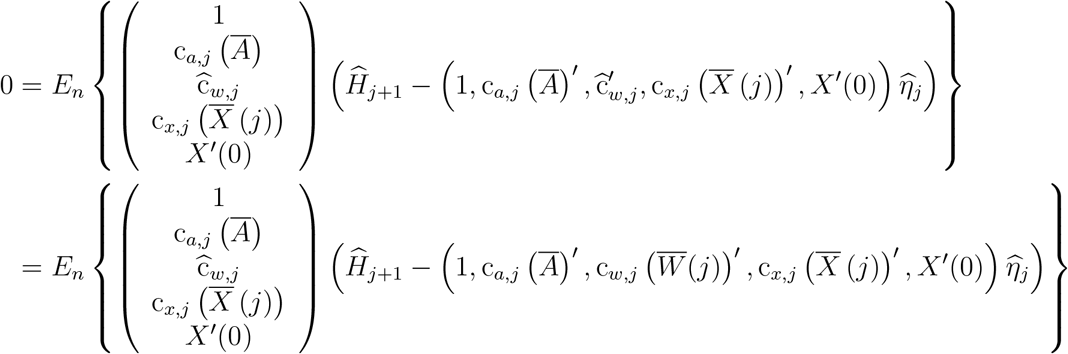

because

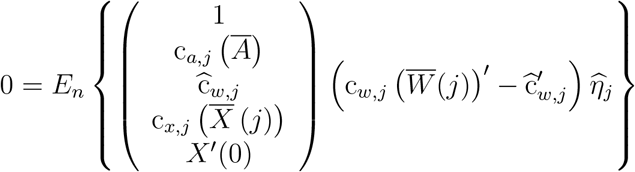

by virtue of 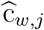 being the least-square projection of 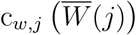 onto

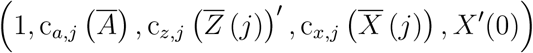

which spans

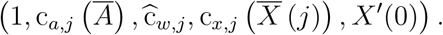

**Table A.1:**
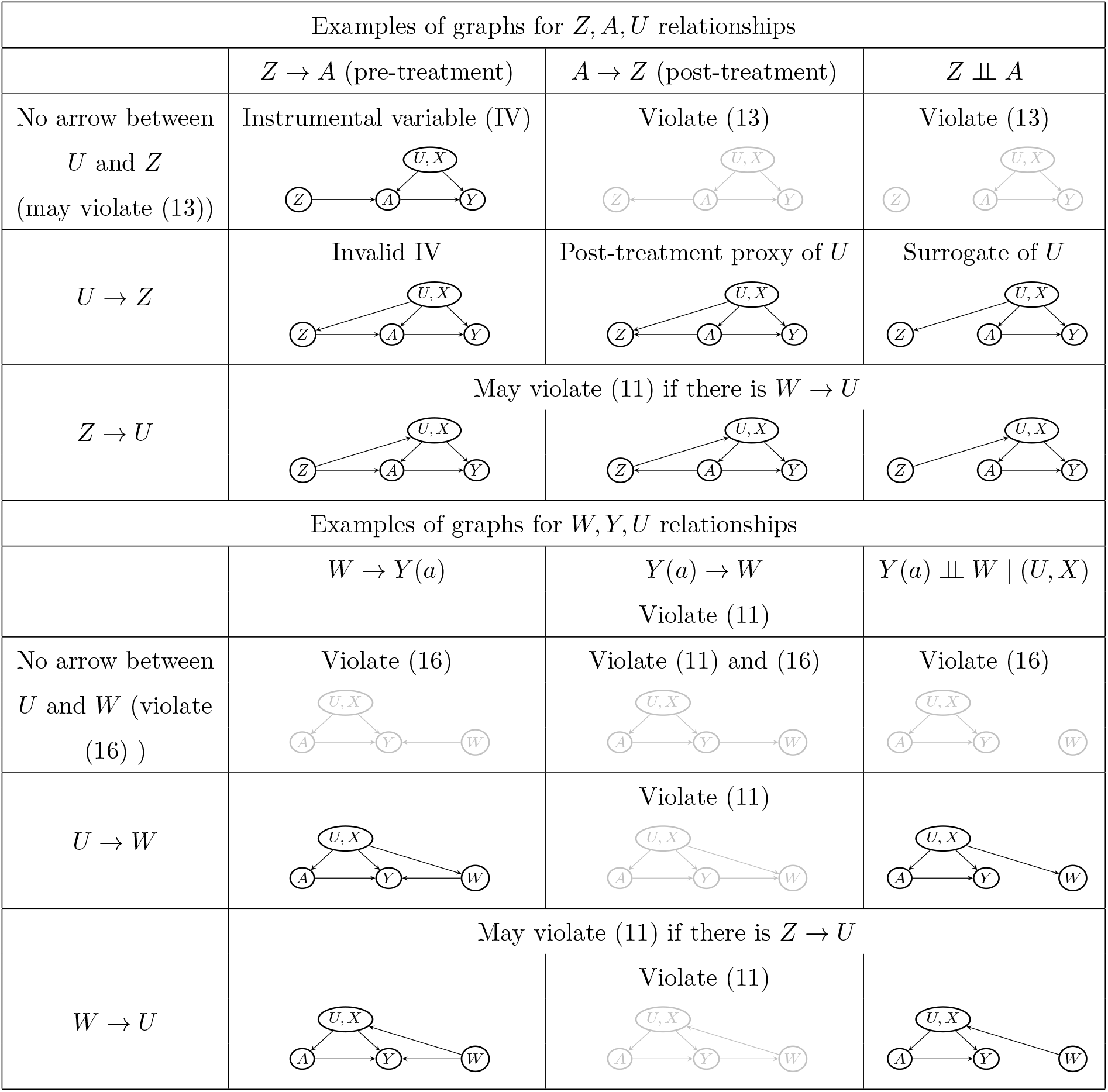
Examples of graphs for *Z, A, U* relationships and for *W, Y, U* relationships. The two pieces of graphs can be combined in to a directed acyclic graph that encodes the assumptions on proxy variables of types b and c. Grey colored graphs are invalid due to violation of key assumptions.

**Table A.2:**
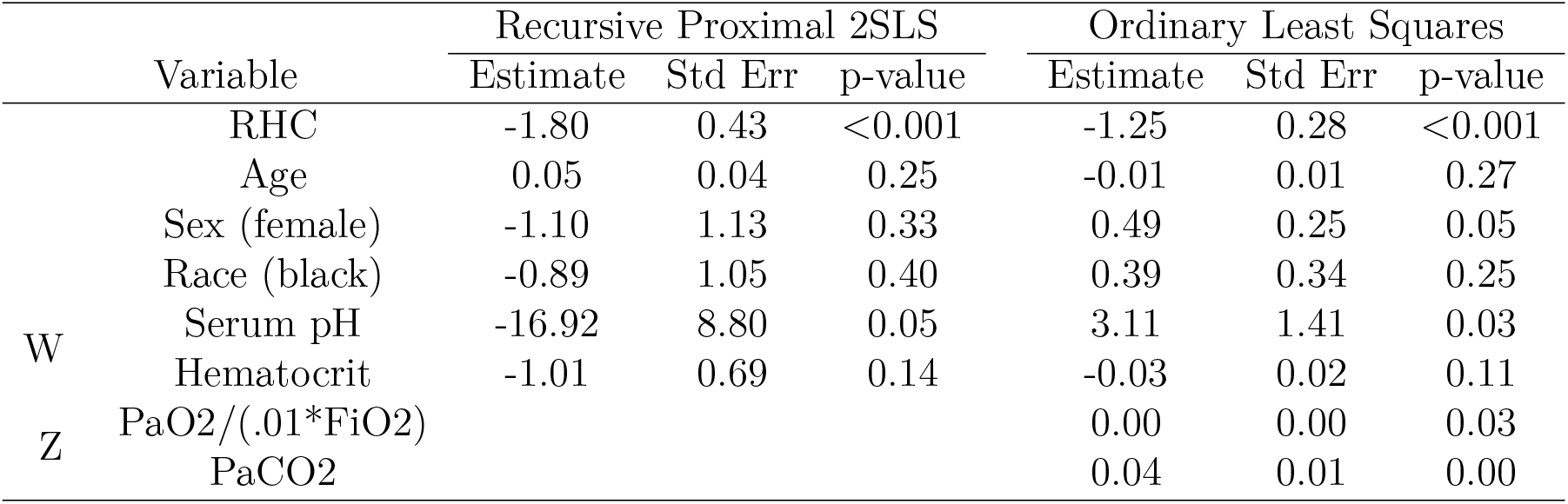
Results from right heart catherization empirical application.

**Table A.3:** Results from Methotrexate empirical application.

| *Dependent variable:* |  |
| --- | --- |
| jc |  |
| mtxspan | -0.154*** (0.057)<br>p = 0.008 |
| fitted_jc | 0.524*** (0.097)<br>p = 0.00000 |
| *Note:* | *p<0.1; **p<0.05; ***p<0.01 |
| h_1 |  |
| cum_treatment | -0.188*** (0.057)<br>p = 0.00000 |
| *Note:* | *p<0.1; **p<0.05; ***p<0.01 |

